# Early detection of SARS-CoV-2 variants using traveler-based genomic surveillance at four US airports, September 2021- January 2022

**DOI:** 10.1101/2022.03.21.22272490

**Authors:** Renee D. Wegrzyn, Grace D. Appiah, Robert Morfino, Scott R. Milford, Allison Taylor Walker, Ezra T. Ernst, William W. Darrow, Siayo Lisa Li, Keith Robison, Duncan MacCannell, Dongjuan Dai, Brintha P. Girinathan, Allison L. Hicks, Bryan Cosca, Gabrielle Woronoff, Alex M. Plocik, Birgitte B. Simen, Leah Moriarty, Sarah Anne J. Guagliardo, Martin S. Cetron, Cindy R. Friedman

## Abstract

We enrolled arriving international air travelers in SARS-CoV-2 genomic surveillance, using molecular testing of pooled nasal swabs, and sequencing positive samples for viral lineage. Traveler-based genomic surveillance provided early warning variant detection; we reported the first U.S. Omicron BA.2 and first BA.3 in North America, weeks before next reported detection.

## Background

Despite layered mitigation measures, international travel during the COVID-19 pandemic continues to facilitate global spread of SARS-CoV-2, including novel variants of concern (VOCs). On November 26, 2021, B.1.1.529 (Omicron) was designated as a VOC by the World Health Organization [1]. On December 6, 2021, as part of measures to reduce the introduction and spread of Omicron, the requirement for a negative SARS-CoV-2 test taken before air travel to the United States was shortened from three days to one day pre-departure [1]. Although SARS-CoV-2 genomic sequencing has increased significantly during the pandemic [2], there is still a gap in early detection of emerging variants among arriving travelers.

In September 2021, the Centers for Disease Control and Prevention (CDC), in collaboration with private partners, implemented a voluntary SARS-CoV-2 genomic surveillance pilot program.

Initially we enrolled arriving air travelers from India. On November 28, we expanded the program to include travelers arriving from countries with high travel volumes, including those where Omicron was first detected.

## Methods

### Design, Setting, and Participants

During September 29–November 27, 2021, the surveillance program included travelers arriving on seven direct flights from India at three international airports: John F. Kennedy, New York (September 29), Newark Liberty, New Jersey (October 4), and San Francisco, California (October 12). During November 28–January 23, Hartsfield-Jackson Atlanta International Airport, Georgia was added, and participation was offered to travelers from South Africa, Nigeria, the United Kingdom, France, Germany, and Brazil, arriving on approximately 50 flights per day. Participants were 18 years or older, provided informed consent, and completed demographic, clinical, and travel history questions.

### Sample Collection

Participants could opt-in for either, in-airport dry nasal swab sample self-collection, at-home saliva sample collection 3-5 days after arrival, or both (Supplementary Figure 1). For in-airport sampling, travelers were asked to self-collect a lower nasal swab sample. Samples were placed in pooled collection tubes with 5–25 other samples, with size adjusted based on flight volume, and shipped to the Concentric Laboratory Network in viral transport media. During September 29–November 27, pooled samples were grouped together based on arriving flight. During November 28-January 23, pooled samples were grouped by country of origin. For the at-home kit, travelers were asked to collect a saliva sample on day 3–5 after arrival and send it to the laboratory.

### Laboratory Testing

All samples underwent SARS-CoV-2 reverse transcription polymerase chain reaction (RT-PCR). After November 27, samples were tested for S-gene target failure (SGTF) using TaqPath COVID-19 assay [3]. All positives underwent whole genome sequencing and variant lineage determination. Reverse transcribed RNA was amplified using the ARTICv3 protocol [4]. Amplicons were pooled and prepared using standard protocols. For Illumina sequencing, samples underwent tagmentation and were sequenced on NovaSeq 6000 (2×50 bp; Illumina). For rapid lineage identification, a ligation-based library was prepared and sequenced on GridION (Oxford Nanopore) as described in supplemental methods.

### Reporting

Travelers participating in pooled testing were advised to submit their at-home kit for individual testing. Individual test results were reported to participants via a secure digital portal and to public health authorities per CDC reporting guidelines [5]. Sequencing data were provided to CDC for viral culture and characterization and uploaded to the Global Initiative on Sharing Avian Influenza Data (GISAID) public database.

### Statistical Analysis

For this analysis we focused on the effectiveness of pooled testing for early variant detection and thus included pooled testing results only. Using Fisher’s exact tests, we assessed differences in pooled positivity rates across two collection periods (September 29–November 27 and November 28–January 23) and by flight country of origin. Statistical analysis was conducted in R 4.0.3. This activity was reviewed by CDC and was conducted consistent with applicable federal law and CDC policy.^1^

## Results

During September 29, 2021–January 23, 2022, we enrolled 16,149 (∼10%) of an estimated 161,000 eligible travelers, yielding 1,454 sample pools. Overall, 221 (16%) of 1,367 pooled samples (average pool size 11 swabs) tested were SARS-CoV-2-positive. The median turnaround time from sample collection to sequencing was 11 business days (range, 5 - 20). For select samples, we performed expedited sequencing within 48 hours to confirm validity of SGTF as an early indicator for Omicron. Positivity increased significantly from 1.8% (6/338) during September 29–November 27 to 20.9% (215/1029) after November 27 (p<0.001). Percent positivity of pooled samples varied significantly by country of flight origin, ranging from 43.5% (40/92) in South Africa, to 9.3% (68/733) in India (p < 0.001) (Supplementary Table 1). Before November 28, all 6 lineages were Delta (B.1.617-like), apart from one undetermined lineage. During November 28–January 23, 67% (145/215) of positive pooled samples collected were Omicron (B.1.1.529-like), 5% (11/215) were Delta (B.1.617-like), and the remaining 27% (59/215) lineages could not be determined due to low sample sequencing coverage (low number of sample nucleotide bases aligned to a reference genome) (Figure 1a and Supplementary Table 2). Of 145 Omicron sequences, 112 exhibited complete or partial SGTF; BA.2 sub-lineages did not. Omicron sub-lineages included BA.1 (100), BA1.1 (12), BA.2 (26), and BA.3 (1), BA.2 + Orf1a:M85 (1), and BA.2 + S:R346K (1). Four samples were identified as Omicron, but sublineage could not be determined due to low sequencing coverage. A sample collected on December 14 was the first reported BA.2 in the United States, 7 days earlier than any other U.S. report (Figure 1b). Similarly, a sample collected on December 3 was the first reported BA.3 in North America, 43 days before the next report [6].

**Figure 1.**
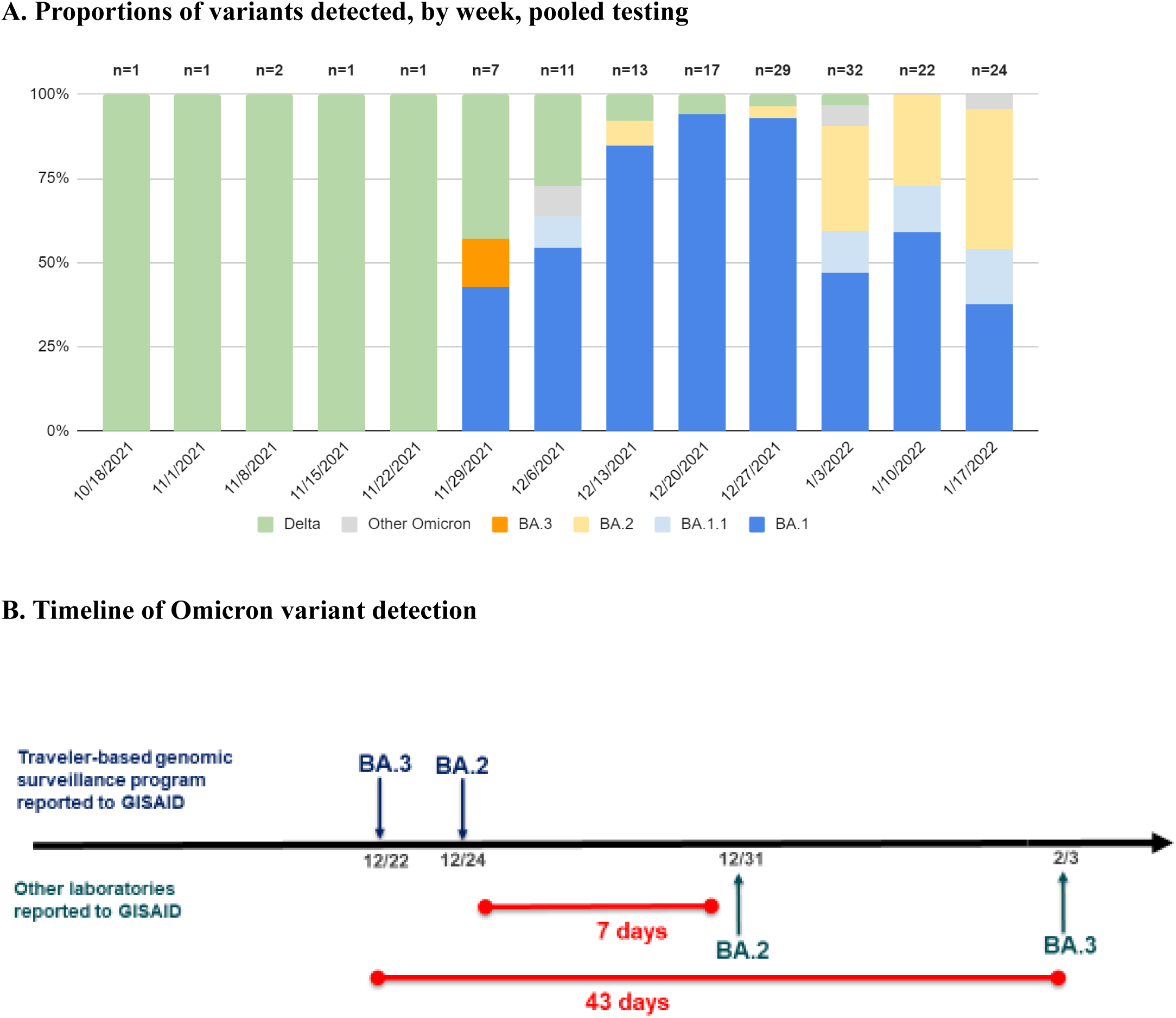
Proportions of variants detected among arriving international travelers and rapid reporting of Omicron subvariants. *A*, Identified variant lineage and sublineage proportions among sequenced samples by collection week, November 28, 2021 to January 23, 2022. *B*, Timeline of reported Omicron subvariants in the United States, comparing first reporting in GISAID of BA.2 and BA.3 by the traveler-based genomic surveillance program (in blue) compared to first reporting from other laboratories (in green). The program’s timeliness of first reporting of BA.2 (on December 24) and BA.3 (on December 22) was 7 and 43 days earlier, respectively, than next reporting in GISAID.

## Discussion

This traveler-based SARS-CoV-2 genomic surveillance program was able to identify importation of variants, including Omicron sub-lineages BA.2 and BA.3 at 7 and 43 days before they were reported elsewhere in the United States and North America, respectively. Overall, 16% of pooled tests were positive, with 21% positivity following Omicron emergence. We detected an increased number of positive post-arrival pooled samples after December 6, when air passengers were required to have a negative sample collected within one day before flight departure.

Possible reasons for high pooled test-positivity on arrival despite negative pre-departure testing include timing of infection (i.e., before infection can be detected by available testing), use of certain testing modalities with lower sensitivity [7], or infection soon after pre-departure sample collection [8, 9]. If passengers had infections that were not yet detected in pre-departure testing, longer flight times may have led to detection of more positive tests on arrival [9]. Many of our positive pools were from flights over 16 hours long. Finally, it is possible that fraudulent test results were presented to meet the pre-departure testing requirement [10].

Pooled testing in this program is advantageous as it enables efficient, large volume sampling and increases testing throughput while conserving resources. This can be valuable for continued detection when prevalence of SARS-CoV-2 infection is low. The pooled testing design minimizes dilution and reduces loss of sensitivity by pooling dry swabs at the point of collection Each Concentric network laboratory is validated to ensure molecular assay sensitivity of 1,500 viral copies/mL. The disadvantage of pooled testing is an inability to directly link test results with individual-level data. Follow-up individual testing, such as the at-home test kits collected in our program (data not presented), provide an additional opportunity to capture linkable meta-data.

With ∼ 10% participation rate, we were able to detect the first Omicron sub-lineages BA.2 and BA.3 not previously reported in the United States or North America, weeks before they were reported by other US and North American sequencing efforts. The country-level proportions of variants that we identified were consistent with those reported by national and global sequencing programs [2]. Our study suggests that when COVID-19 rates are high, as during the Omicron surge, a 10% participation rate would be sufficient to detect relatively rare variant sublineages. As COVID-19 rates decline in the post-Omicron period, the program could shift to population-level genomic surveillance techniques that are not reliant on passenger participation such as wastewater or air sampling from aircraft environments to increase variant detection.

Detection of imported emerging infectious diseases has traditionally focused on travelers presenting to health clinics after symptom onset [12]. COVID-19 presents unique challenges since transmission often occurs before symptom onset or in asymptomatic infected persons [7]. By the time variants are detected there is often widespread community transmission. Many countries have required testing for arriving travelers to limit introduction and spread of SARS-CoV-2 [11] yet few utilize traveler-based viral genomic surveillance to detect new variants and provide more detailed epidemiological data. Earlier detection of new SARS-CoV-2 variants of interest and concern allows researchers and public health officials the needed time to gather information about transmissibility, virulence, and vaccine effectiveness to enable adjustments to treatment and prevention strategies [2].

This traveler-based genomic surveillance program underscores the importance of public-private partnerships in achieving public health priorities in an ever-changing pandemic, and the utility of surveillance tools beyond traditional individual testing. The program’s scalability and adaptability, including the ability to rapidly add locations and expedite sequencing, were key factors for success. Traveler-based SARS-CoV-2 genomic surveillance provides a model of pathogen detection that can be used as an early warning, sentinel system for future outbreaks affecting international travelers or pandemics.

## Data Availability

All data produced in the present study are available upon reasonable request to the authors

## Funding for this manuscript

This work was supported by the Centers for Disease Control and Prevention contract award number 75D30121C12036.

## Acknowledgements

The authors would like to thank the entire XpresCheck (Erica Mares, Lesley Shirley, Rob Stein, Henry Streich, Miguel Yapor), Concentric by Ginkgo (Thomas Aichele, Dan Bayley, Juskarun Cheema, Tyler Clarkson, Aakash Desai, Joseph Fridman, Alix Hamilton, Corey Hoehn, Erica Jackson, Hannah Knoll, Frank Langston, John Mcbride, Justin Montgomery, Jason Ng, Rich Nordin, Sarah Rush, Zach Smith, Sativa Turner, Erika Gute, Maria Pis-Lopez, Cherish Weiler), the CDC Division of Global Migration and Quarantine Travelers’ Health (Teresa Smith, Jessica Allen, Laura Leidel, Igor Ristic, Robin Rinker) and Quarantine and Border Health Services Branches (Clive Brown, Tai-Ho Chen, Alida Gertz, Matthew Palo), CDC COVID-19 Response Health Department Liaisons, US Customs and Border Protection, and state and local public health authorities - who supported the operational implementation of the program on the ground in airports.

## Disclaimer

The findings and conclusions of this report are those of the authors and do not necessarily represent the official position of the Centers for Disease Control and Prevention. This activity was conducted in partnership with commercial partners, Xprescheck and Concentric by Ginkgo. The use of products’ or services’ names is for identification purposes and does not mean endorsement by the Centers for Disease Control and Prevention.

## Supplementary Data

**Supplementary Figure 1.**
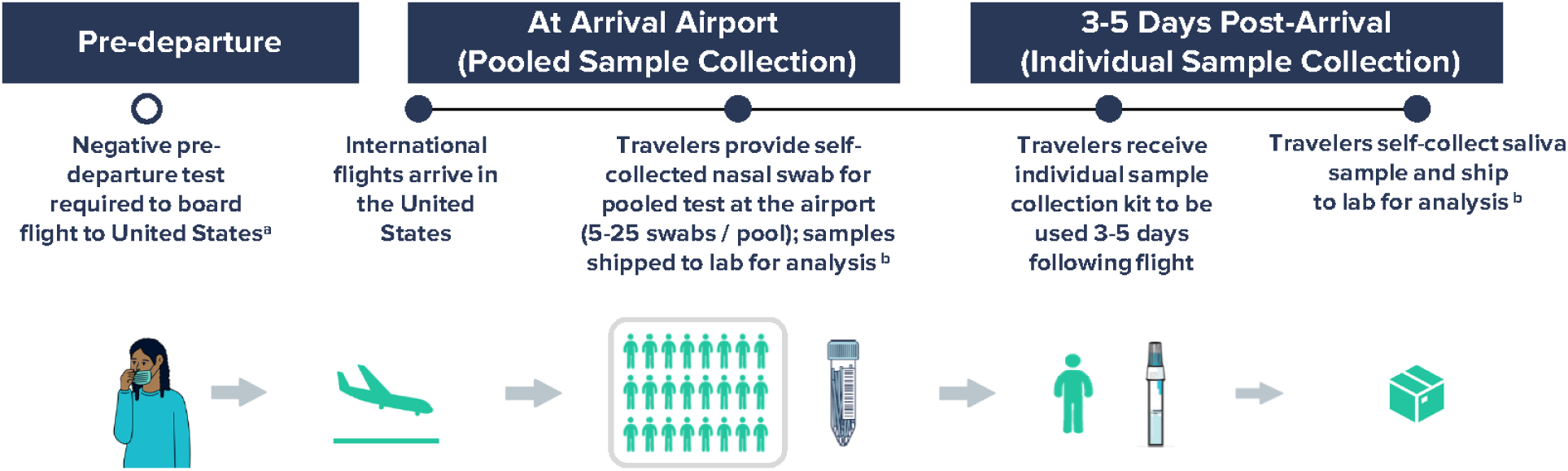
Workflow for airport pooled and individual sample collection to detect SARS-CoV-2 variants of concern as part of an international arriving traveler surveillance program, United States, September 29 – January 23, 2022. ^a^ Negative COVID-19 test required to board flight to the United States per CDC Order; the time window for pre-departure testing was shortened from 3 days to 1 day on December 6, 2021. ^b^ Analyses included RT-PCR testing of all samples within 24-48 hours of sample collection and whole genome sequencing of all positive samples in a median of 11 days.

**Supplementary Table 1.** Results of viral genomic sequencing to detect SARS-CoV-2 variants among arriving international travelers at four US airports by flight origin country, September 29 – January 23, 2022.

| Flight Origin | Participants | SARS-CoV-2<br>Positive/Tested (%)* | Omicron<br>Omicron/ SARS-CoV-2 Positive<br>Pools (%) |
| --- | --- | --- | --- |
| South Africa | 1,129 | 40/92 (43.5) | 31/40 (77.5) |
| Brazil | 529 | 15/46 (32.6) | 10/15 (66.7) |
| France | 1,274 | 30/120 (25.0) | 18/30 (60.0) |
| United Kingdom | 1,627 | 30/162 (18.5) | 15/30 (50.0) |
| Germany | 2,325 | 38/213 (17.8) | 23/38 (60.5) |
| India | 9,197 | 68/732 (9.3) | 46/68 (67.6) |
| N/A | 68 | 0/2 (0.0) | 0/0 N/A |
| **Total** | **16,149** | **221/1367 (16.1)** | **143/221 (64.7)** |

\*Fisher's exact test p-value <0.001.

**Supplementary Table 2.**
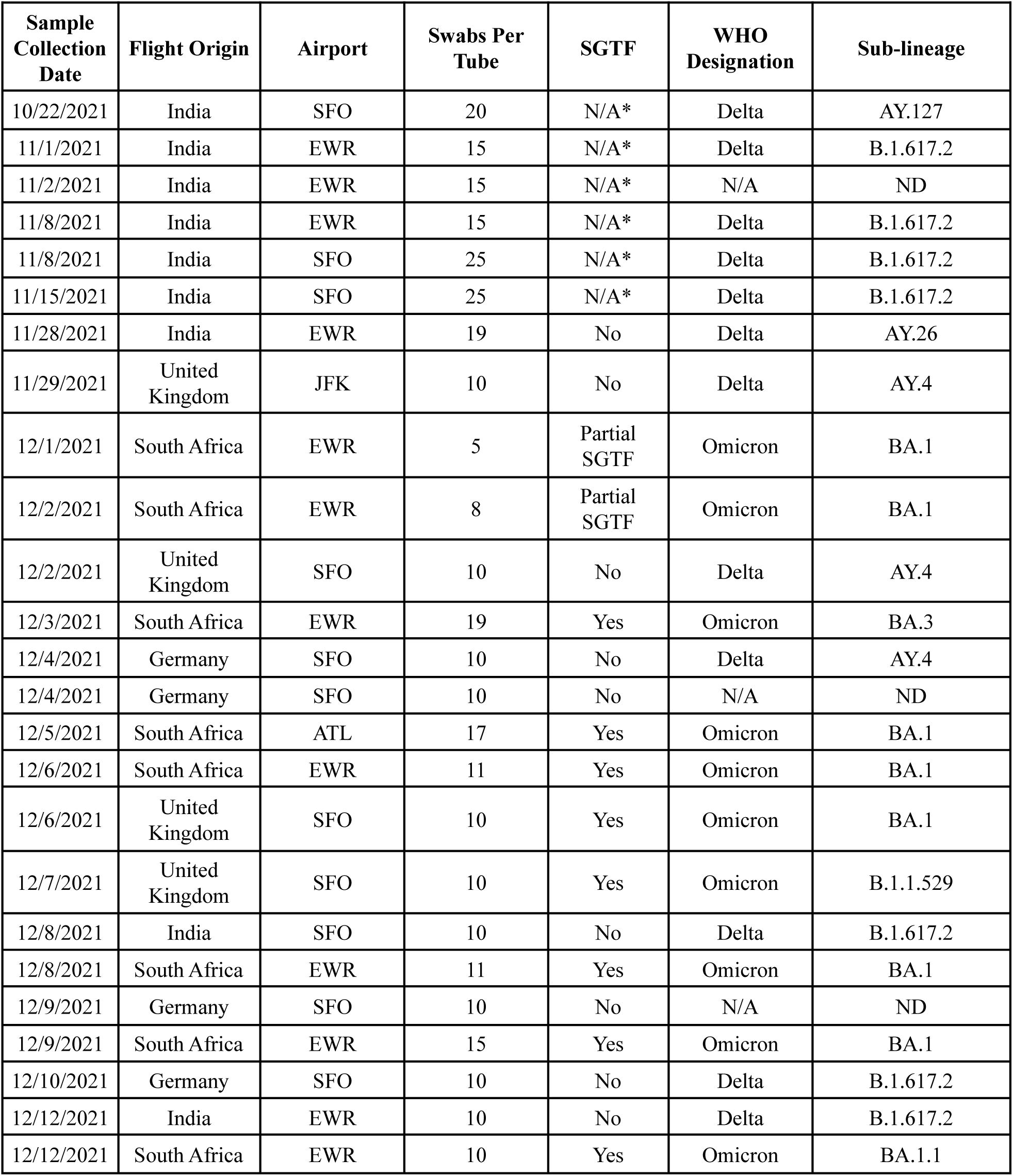
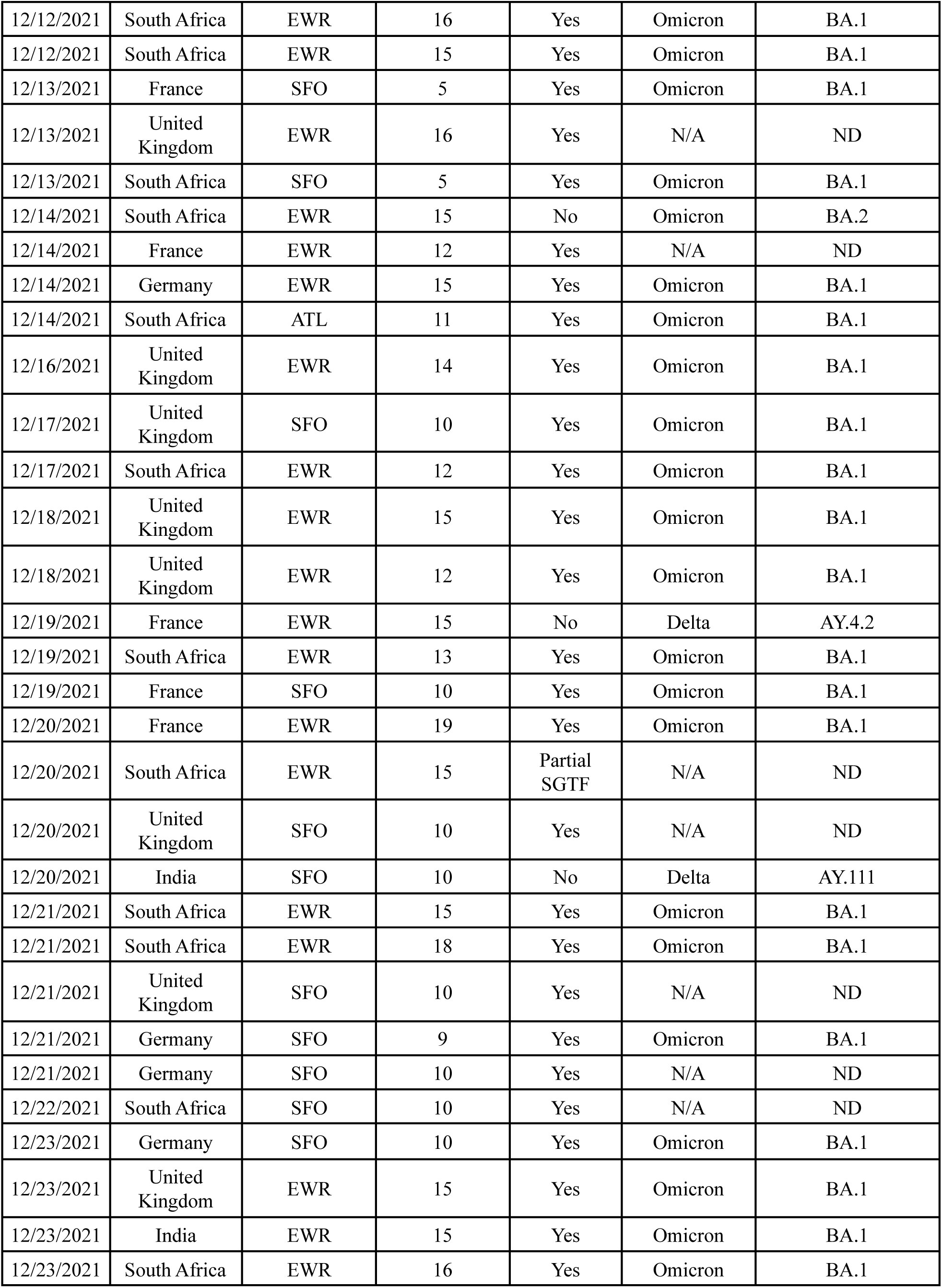
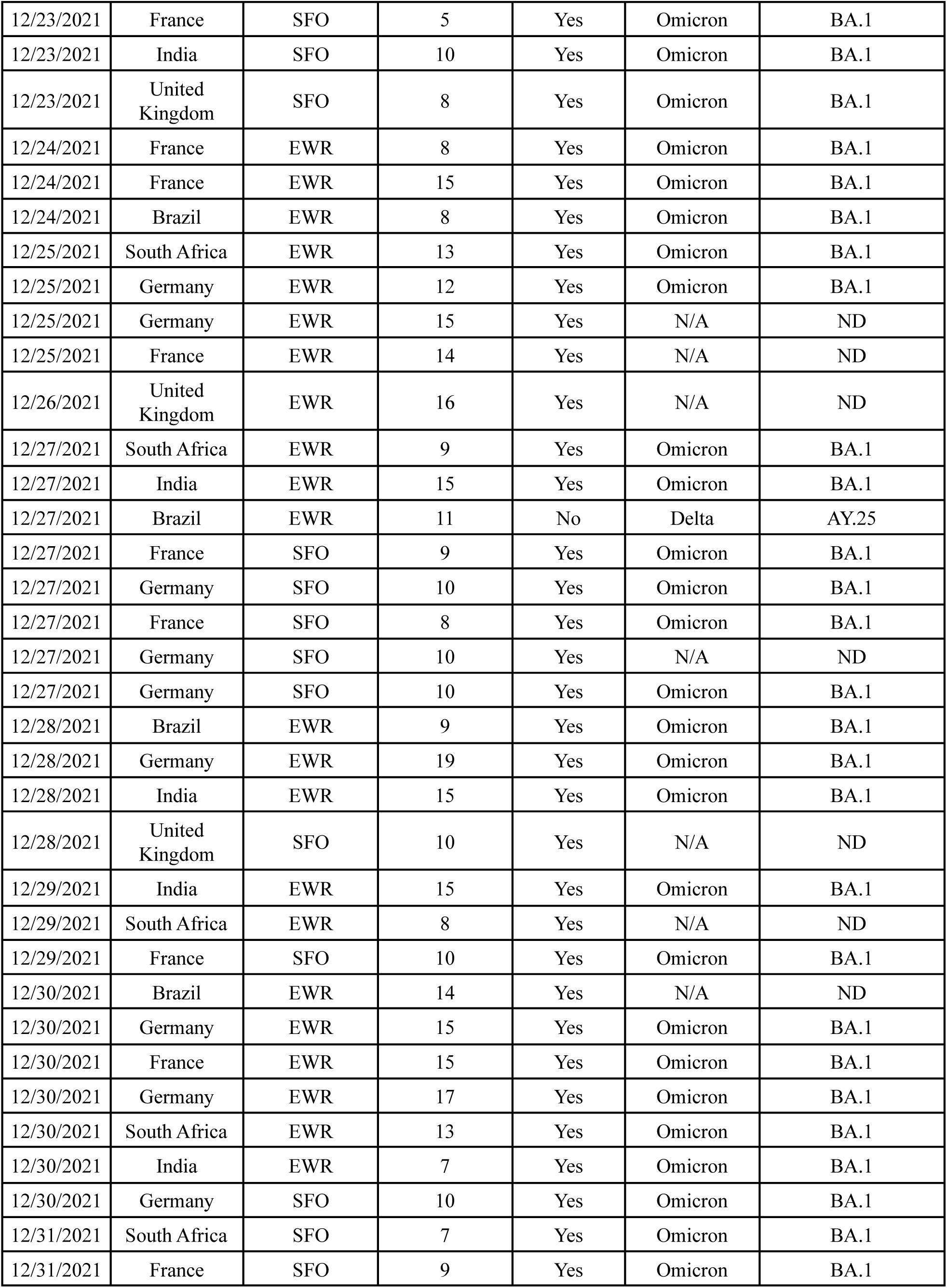
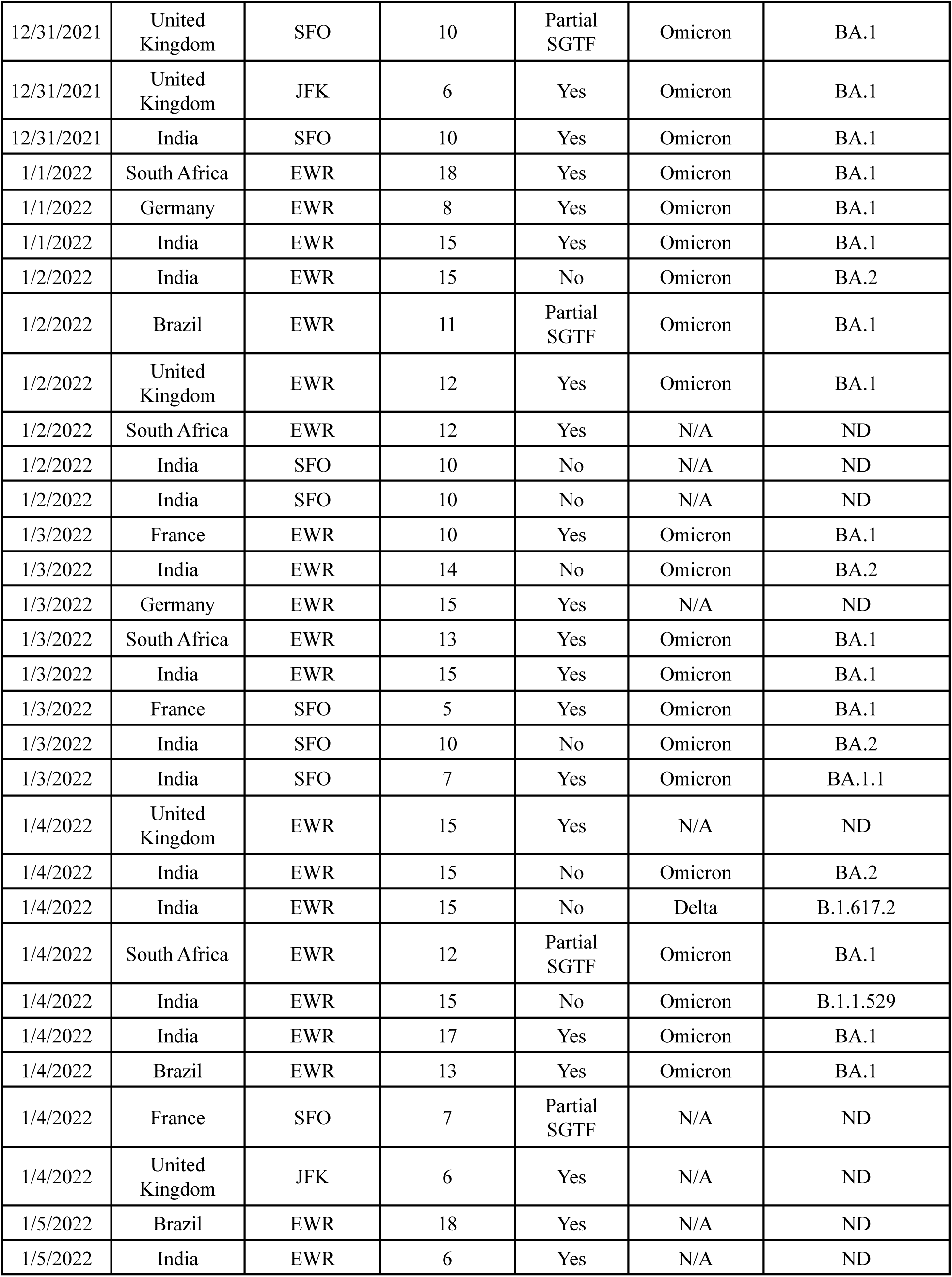
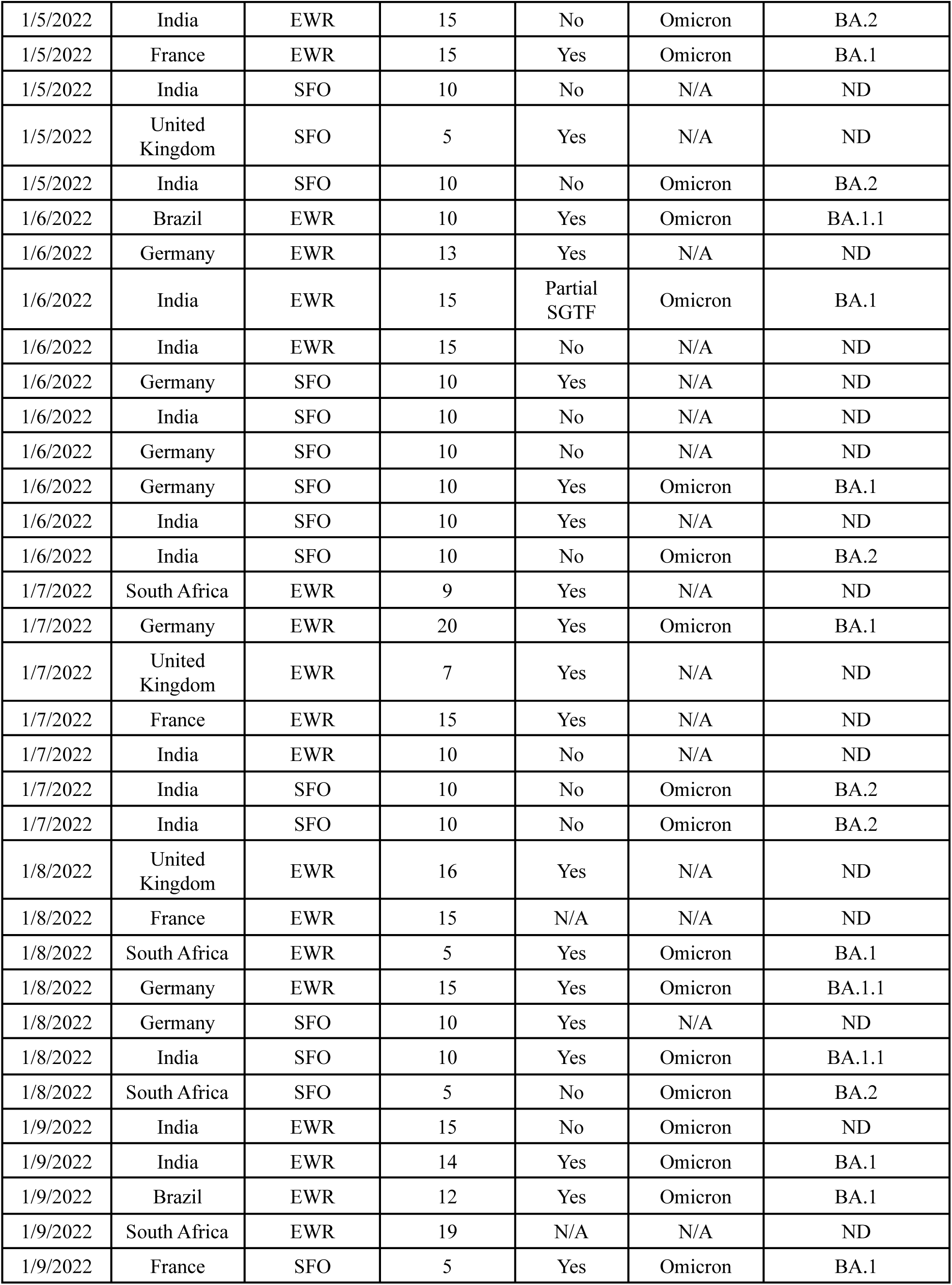
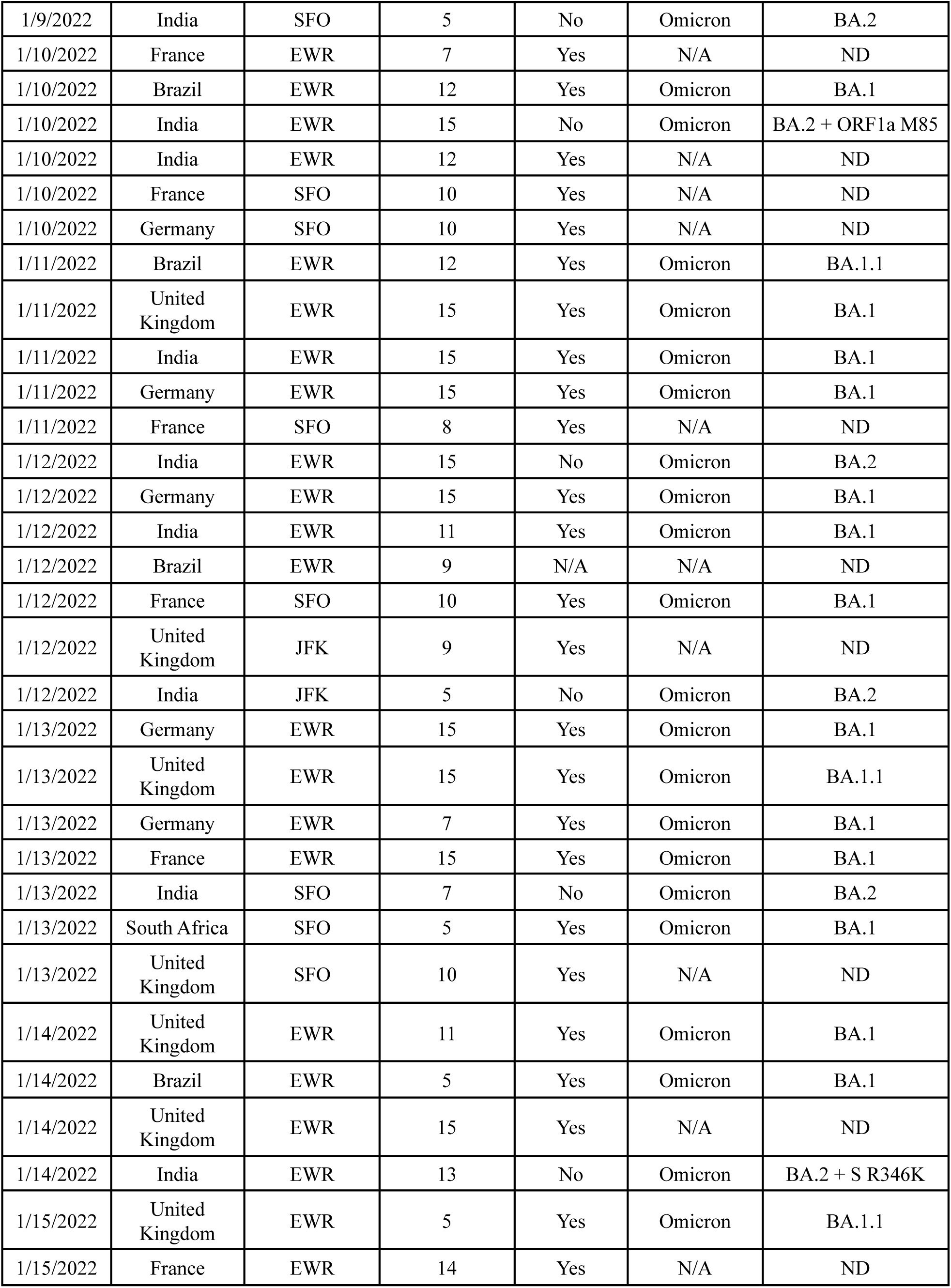
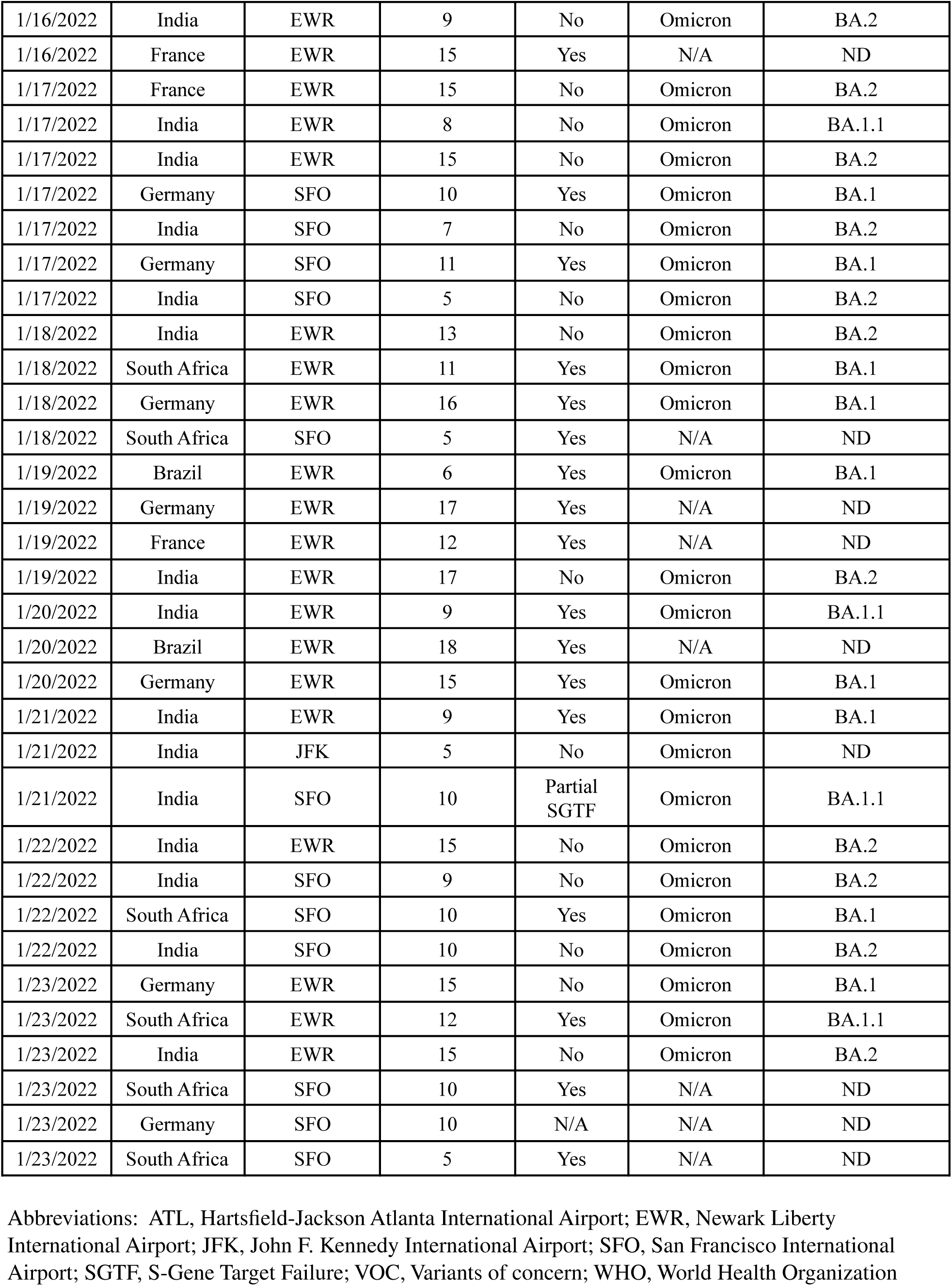
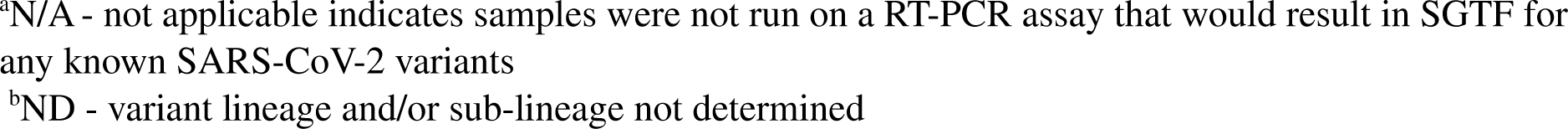
Whole genome sequencing results of 222 SARS-CoV-2-positive pooled samples collected from arriving international travelers at four US airports, September 29 – January 23, 2022.

| Sample Collection Date | Flight Origin | Airport | Swabs Per Tube | SGTF | WHO Designation | Sub-lineage |
| --- | --- | --- | --- | --- | --- | --- |
| 10/22/2021 | India | SFO | 20 | N/A* | Delta | AY.127 |
| 11/1/2021 | India | EWR | 15 | N/A* | Delta | B.1.617.2 |
| 11/2/2021 | India | EWR | 15 | N/A* | N/A | ND |
| 11/8/2021 | India | EWR | 15 | N/A* | Delta | B.1.617.2 |
| 11/8/2021 | India | SFO | 25 | N/A* | Delta | B.1.617.2 |
| 11/15/2021 | India | SFO | 25 | N/A* | Delta | B.1.617.2 |
| 11/28/2021 | India | EWR | 19 | No | Delta | AY.26 |
| 11/29/2021 | United Kingdom | JFK | 10 | No | Delta | AY.4 |
| 12/1/2021 | South Africa | EWR | 5 | Partial SGTF | Omicron | BA.1 |
| 12/2/2021 | South Africa | EWR | 8 | Partial SGTF | Omicron | BA.1 |
| 12/2/2021 | United Kingdom | SFO | 10 | No | Delta | AY.4 |
| 12/3/2021 | South Africa | EWR | 19 | Yes | Omicron | BA.3 |
| 12/4/2021 | Germany | SFO | 10 | No | Delta | AY.4 |
| 12/4/2021 | Germany | SFO | 10 | No | N/A | ND |
| 12/5/2021 | South Africa | ATL | 17 | Yes | Omicron | BA.1 |
| 12/6/2021 | South Africa | EWR | 11 | Yes | Omicron | BA.1 |
| 12/6/2021 | United Kingdom | SFO | 10 | Yes | Omicron | BA.1 |
| 12/7/2021 | United Kingdom | SFO | 10 | Yes | Omicron | B.1.1.529 |
| 12/8/2021 | India | SFO | 10 | No | Delta | B.1.617.2 |
| 12/8/2021 | South Africa | EWR | 11 | Yes | Omicron | BA.1 |
| 12/9/2021 | Germany | SFO | 10 | No | N/A | ND |
| 12/9/2021 | South Africa | EWR | 15 | Yes | Omicron | BA.1 |
| 12/10/2021 | Germany | SFO | 10 | No | Delta | B.1.617.2 |
| 12/12/2021 | India | EWR | 10 | No | Delta | B.1.617.2 |
| 12/12/2021 | South Africa | EWR | 10 | Yes | Omicron | BA.1.1 |
| 12/12/2021 | South Africa | EWR | 16 | Yes | Omicron | BA.1 |
| 12/12/2021 | South Africa | EWR | 15 | Yes | Omicron | BA.1 |
| 12/13/2021 | France | SFO | 5 | Yes | Omicron | BA.1 |
| 12/13/2021 | United Kingdom | EWR | 16 | Yes | N/A | ND |
| 12/13/2021 | South Africa | SFO | 5 | Yes | Omicron | BA.1 |
| 12/14/2021 | South Africa | EWR | 15 | No | Omicron | BA.2 |
| 12/14/2021 | France | EWR | 12 | Yes | N/A | ND |
| 12/14/2021 | Germany | EWR | 15 | Yes | Omicron | BA.1 |
| 12/14/2021 | South Africa | ATL | 11 | Yes | Omicron | BA.1 |
| 12/16/2021 | United Kingdom | EWR | 14 | Yes | Omicron | BA.1 |
| 12/17/2021 | United Kingdom | SFO | 10 | Yes | Omicron | BA.1 |
| 12/17/2021 | South Africa | EWR | 12 | Yes | Omicron | BA.1 |
| 12/18/2021 | United Kingdom | EWR | 15 | Yes | Omicron | BA.1 |
| 12/18/2021 | United Kingdom | EWR | 12 | Yes | Omicron | BA.1 |
| 12/19/2021 | France | EWR | 15 | No | Delta | AY.4.2 |
| 12/19/2021 | South Africa | EWR | 13 | Yes | Omicron | BA.1 |
| 12/19/2021 | France | SFO | 10 | Yes | Omicron | BA.1 |
| 12/20/2021 | France | EWR | 19 | Yes | Omicron | BA.1 |
| 12/20/2021 | South Africa | EWR | 15 | Partial SGTF | N/A | ND |
| 12/20/2021 | United Kingdom | SFO | 10 | Yes | N/A | ND |
| 12/20/2021 | India | SFO | 10 | No | Delta | AY.111 |
| 12/21/2021 | South Africa | EWR | 15 | Yes | Omicron | BA.1 |
| 12/21/2021 | South Africa | EWR | 18 | Yes | Omicron | BA.1 |
| 12/21/2021 | United Kingdom | SFO | 10 | Yes | N/A | ND |
| 12/21/2021 | Germany | SFO | 9 | Yes | Omicron | BA.1 |
| 12/21/2021 | Germany | SFO | 10 | Yes | N/A | ND |
| 12/22/2021 | South Africa | SFO | 10 | Yes | N/A | ND |
| 12/23/2021 | Germany | SFO | 10 | Yes | Omicron | BA.1 |
| 12/23/2021 | United Kingdom | EWR | 15 | Yes | Omicron | BA.1 |
| 12/23/2021 | India | EWR | 15 | Yes | Omicron | BA.1 |
| 12/23/2021 | South Africa | EWR | 16 | Yes | Omicron | BA.1 |
| 12/23/2021 | France | SFO | 5 | Yes | Omicron | BA.1 |
| 12/23/2021 | India | SFO | 10 | Yes | Omicron | BA.1 |
| 12/23/2021 | United Kingdom | SFO | 8 | Yes | Omicron | BA.1 |
| 12/24/2021 | France | EWR | 8 | Yes | Omicron | BA.1 |
| 12/24/2021 | France | EWR | 15 | Yes | Omicron | BA.1 |
| 12/24/2021 | Brazil | EWR | 8 | Yes | Omicron | BA.1 |
| 12/25/2021 | South Africa | EWR | 13 | Yes | Omicron | BA.1 |
| 12/25/2021 | Germany | EWR | 12 | Yes | Omicron | BA.1 |
| 12/25/2021 | Germany | EWR | 15 | Yes | N/A | ND |
| 12/25/2021 | France | EWR | 14 | Yes | N/A | ND |
| 12/26/2021 | United Kingdom | EWR | 16 | Yes | N/A | ND |
| 12/27/2021 | South Africa | EWR | 9 | Yes | Omicron | BA.1 |
| 12/27/2021 | India | EWR | 15 | Yes | Omicron | BA.1 |
| 12/27/2021 | Brazil | EWR | 11 | No | Delta | AY.25 |
| 12/27/2021 | France | SFO | 9 | Yes | Omicron | BA.1 |
| 12/27/2021 | Germany | SFO | 10 | Yes | Omicron | BA.1 |
| 12/27/2021 | France | SFO | 8 | Yes | Omicron | BA.1 |
| 12/27/2021 | Germany | SFO | 10 | Yes | N/A | ND |
| 12/27/2021 | Germany | SFO | 10 | Yes | Omicron | BA.1 |
| 12/28/2021 | Brazil | EWR | 9 | Yes | Omicron | BA.1 |
| 12/28/2021 | Germany | EWR | 19 | Yes | Omicron | BA.1 |
| 12/28/2021 | India | EWR | 15 | Yes | Omicron | BA.1 |
| 12/28/2021 | United Kingdom | SFO | 10 | Yes | N/A | ND |
| 12/29/2021 | India | EWR | 15 | Yes | Omicron | BA.1 |
| 12/29/2021 | South Africa | EWR | 8 | Yes | N/A | ND |
| 12/29/2021 | France | SFO | 10 | Yes | Omicron | BA.1 |
| 12/30/2021 | Brazil | EWR | 14 | Yes | N/A | ND |
| 12/30/2021 | Germany | EWR | 15 | Yes | Omicron | BA.1 |
| 12/30/2021 | France | EWR | 15 | Yes | Omicron | BA.1 |
| 12/30/2021 | Germany | EWR | 17 | Yes | Omicron | BA.1 |
| 12/30/2021 | South Africa | EWR | 13 | Yes | Omicron | BA.1 |
| 12/30/2021 | India | EWR | 7 | Yes | Omicron | BA.1 |
| 12/30/2021 | Germany | SFO | 10 | Yes | Omicron | BA.1 |
| 12/31/2021 | South Africa | SFO | 7 | Yes | Omicron | BA.1 |
| 12/31/2021 | France | SFO | 9 | Yes | Omicron | BA.1 |
| 12/31/2021 | United Kingdom | SFO | 10 | Partial SGTF | Omicron | BA.1 |
| 12/31/2021 | United Kingdom | JFK | 6 | Yes | Omicron | BA.1 |
| 12/31/2021 | India | SFO | 10 | Yes | Omicron | BA.1 |
| 1/1/2022 | South Africa | EWR | 18 | Yes | Omicron | BA.1 |
| 1/1/2022 | Germany | EWR | 8 | Yes | Omicron | BA.1 |
| 1/1/2022 | India | EWR | 15 | Yes | Omicron | BA.1 |
| 1/2/2022 | India | EWR | 15 | No | Omicron | BA.2 |
| 1/2/2022 | Brazil | EWR | 11 | Partial SGTF | Omicron | BA.1 |
| 1/2/2022 | United Kingdom | EWR | 12 | Yes | Omicron | BA.1 |
| 1/2/2022 | South Africa | EWR | 12 | Yes | N/A | ND |
| 1/2/2022 | India | SFO | 10 | No | N/A | ND |
| 1/2/2022 | India | SFO | 10 | No | N/A | ND |
| 1/3/2022 | France | EWR | 10 | Yes | Omicron | BA.1 |
| 1/3/2022 | India | EWR | 14 | No | Omicron | BA.2 |
| 1/3/2022 | Germany | EWR | 15 | Yes | N/A | ND |
| 1/3/2022 | South Africa | EWR | 13 | Yes | Omicron | BA.1 |
| 1/3/2022 | India | EWR | 15 | Yes | Omicron | BA.1 |
| 1/3/2022 | France | SFO | 5 | Yes | Omicron | BA.1 |
| 1/3/2022 | India | SFO | 10 | No | Omicron | BA.2 |
| 1/3/2022 | India | SFO | 7 | Yes | Omicron | BA.1.1 |
| 1/4/2022 | United Kingdom | EWR | 15 | Yes | N/A | ND |
| 1/4/2022 | India | EWR | 15 | No | Omicron | BA.2 |
| 1/4/2022 | India | EWR | 15 | No | Delta | B.1.617.2 |
| 1/4/2022 | South Africa | EWR | 12 | Partial SGTF | Omicron | BA.1 |
| 1/4/2022 | India | EWR | 15 | No | Omicron | B.1.1.529 |
| 1/4/2022 | India | EWR | 17 | Yes | Omicron | BA.1 |
| 1/4/2022 | Brazil | EWR | 13 | Yes | Omicron | BA.1 |
| 1/4/2022 | France | SFO | 7 | Partial SGTF | N/A | ND |
| 1/4/2022 | United Kingdom | JFK | 6 | Yes | N/A | ND |
| 1/5/2022 | Brazil | EWR | 18 | Yes | N/A | ND |
| 1/5/2022 | India | EWR | 6 | Yes | N/A | ND |
| 1/5/2022 | India | EWR | 15 | No | Omicron | BA.2 |
| 1/5/2022 | France | EWR | 15 | Yes | Omicron | BA.1 |
| 1/5/2022 | India | SFO | 10 | No | N/A | ND |
| 1/5/2022 | United Kingdom | SFO | 5 | Yes | N/A | ND |
| 1/5/2022 | India | SFO | 10 | No | Omicron | BA.2 |
| 1/6/2022 | Brazil | EWR | 10 | Yes | Omicron | BA.1.1 |
| 1/6/2022 | Germany | EWR | 13 | Yes | N/A | ND |
| 1/6/2022 | India | EWR | 15 | Partial SGTF | Omicron | BA.1 |
| 1/6/2022 | India | EWR | 15 | No | N/A | ND |
| 1/6/2022 | Germany | SFO | 10 | Yes | N/A | ND |
| 1/6/2022 | India | SFO | 10 | No | N/A | ND |
| 1/6/2022 | Germany | SFO | 10 | No | N/A | ND |
| 1/6/2022 | Germany | SFO | 10 | Yes | Omicron | BA.1 |
| 1/6/2022 | India | SFO | 10 | Yes | N/A | ND |
| 1/6/2022 | India | SFO | 10 | No | Omicron | BA.2 |
| 1/7/2022 | South Africa | EWR | 9 | Yes | N/A | ND |
| 1/7/2022 | Germany | EWR | 20 | Yes | Omicron | BA.1 |
| 1/7/2022 | United Kingdom | EWR | 7 | Yes | N/A | ND |
| 1/7/2022 | France | EWR | 15 | Yes | N/A | ND |
| 1/7/2022 | India | EWR | 10 | No | N/A | ND |
| 1/7/2022 | India | SFO | 10 | No | Omicron | BA.2 |
| 1/7/2022 | India | SFO | 10 | No | Omicron | BA.2 |
| 1/8/2022 | United Kingdom | EWR | 16 | Yes | N/A | ND |
| 1/8/2022 | France | EWR | 15 | N/A | N/A | ND |
| 1/8/2022 | South Africa | EWR | 5 | Yes | Omicron | BA.1 |
| 1/8/2022 | Germany | EWR | 15 | Yes | Omicron | BA.1.1 |
| 1/8/2022 | Germany | SFO | 10 | Yes | N/A | ND |
| 1/8/2022 | India | SFO | 10 | Yes | Omicron | BA.1.1 |
| 1/8/2022 | South Africa | SFO | 5 | No | Omicron | BA.2 |
| 1/9/2022 | India | EWR | 15 | No | Omicron | ND |
| 1/9/2022 | India | EWR | 14 | Yes | Omicron | BA.1 |
| 1/9/2022 | Brazil | EWR | 12 | Yes | Omicron | BA.1 |
| 1/9/2022 | South Africa | EWR | 19 | N/A | N/A | ND |
| 1/9/2022 | France | SFO | 5 | Yes | Omicron | BA.1 |
| 1/9/2022 | India | SFO | 5 | No | Omicron | BA.2 |
| 1/10/2022 | France | EWR | 7 | Yes | N/A | ND |
| 1/10/2022 | Brazil | EWR | 12 | Yes | Omicron | BA.1 |
| 1/10/2022 | India | EWR | 15 | No | Omicron | BA.2 + ORF1a M85 |
| 1/10/2022 | India | EWR | 12 | Yes | N/A | ND |
| 1/10/2022 | France | SFO | 10 | Yes | N/A | ND |
| 1/10/2022 | Germany | SFO | 10 | Yes | N/A | ND |
| 1/11/2022 | Brazil | EWR | 12 | Yes | Omicron | BA.1.1 |
| 1/11/2022 | United Kingdom | EWR | 15 | Yes | Omicron | BA.1 |
| 1/11/2022 | India | EWR | 15 | Yes | Omicron | BA.1 |
| 1/11/2022 | Germany | EWR | 15 | Yes | Omicron | BA.1 |
| 1/11/2022 | France | SFO | 8 | Yes | N/A | ND |
| 1/12/2022 | India | EWR | 15 | No | Omicron | BA.2 |
| 1/12/2022 | Germany | EWR | 15 | Yes | Omicron | BA.1 |
| 1/12/2022 | India | EWR | 11 | Yes | Omicron | BA.1 |
| 1/12/2022 | Brazil | EWR | 9 | N/A | N/A | ND |
| 1/12/2022 | France | SFO | 10 | Yes | Omicron | BA.1 |
| 1/12/2022 | United Kingdom | JFK | 9 | Yes | N/A | ND |
| 1/12/2022 | India | JFK | 5 | No | Omicron | BA.2 |
| 1/13/2022 | Germany | EWR | 15 | Yes | Omicron | BA.1 |
| 1/13/2022 | United Kingdom | EWR | 15 | Yes | Omicron | BA.1.1 |
| 1/13/2022 | Germany | EWR | 7 | Yes | Omicron | BA.1 |
| 1/13/2022 | France | EWR | 15 | Yes | Omicron | BA.1 |
| 1/13/2022 | India | SFO | 7 | No | Omicron | BA.2 |
| 1/13/2022 | South Africa | SFO | 5 | Yes | Omicron | BA.1 |
| 1/13/2022 | United Kingdom | SFO | 10 | Yes | N/A | ND |
| 1/14/2022 | United Kingdom | EWR | 11 | Yes | Omicron | BA.1 |
| 1/14/2022 | Brazil | EWR | 5 | Yes | Omicron | BA.1 |
| 1/14/2022 | United Kingdom | EWR | 15 | Yes | N/A | ND |
| 1/14/2022 | India | EWR | 13 | No | Omicron | BA.2 + S R346K |
| 1/15/2022 | United Kingdom | EWR | 5 | Yes | Omicron | BA.1.1 |
| 1/15/2022 | France | EWR | 14 | Yes | N/A | ND |
| 1/16/2022 | India | EWR | 9 | No | Omicron | BA.2 |
| 1/16/2022 | France | EWR | 15 | Yes | N/A | ND |
| 1/17/2022 | France | EWR | 15 | No | Omicron | BA.2 |
| 1/17/2022 | India | EWR | 8 | No | Omicron | BA.1.1 |
| 1/17/2022 | India | EWR | 15 | No | Omicron | BA.2 |
| 1/17/2022 | Germany | SFO | 10 | Yes | Omicron | BA.1 |
| 1/17/2022 | India | SFO | 7 | No | Omicron | BA.2 |
| 1/17/2022 | Germany | SFO | 11 | Yes | Omicron | BA.1 |
| 1/17/2022 | India | SFO | 5 | No | Omicron | BA.2 |
| 1/18/2022 | India | EWR | 13 | No | Omicron | BA.2 |
| 1/18/2022 | South Africa | EWR | 11 | Yes | Omicron | BA.1 |
| 1/18/2022 | Germany | EWR | 16 | Yes | Omicron | BA.1 |
| 1/18/2022 | South Africa | SFO | 5 | Yes | N/A | ND |
| 1/19/2022 | Brazil | EWR | 6 | Yes | Omicron | BA.1 |
| 1/19/2022 | Germany | EWR | 17 | Yes | N/A | ND |
| 1/19/2022 | France | EWR | 12 | Yes | N/A | ND |
| 1/19/2022 | India | EWR | 17 | No | Omicron | BA.2 |
| 1/20/2022 | India | EWR | 9 | Yes | Omicron | BA.1.1 |
| 1/20/2022 | Brazil | EWR | 18 | Yes | N/A | ND |
| 1/20/2022 | Germany | EWR | 15 | Yes | Omicron | BA.1 |
| 1/21/2022 | India | EWR | 9 | Yes | Omicron | BA.1 |
| 1/21/2022 | India | JFK | 5 | No | Omicron | ND |
| 1/21/2022 | India | SFO | 10 | Partial<br>SGTF | Omicron | BA.1.1 |
| 1/22/2022 | India | EWR | 15 | No | Omicron | BA.2 |
| 1/22/2022 | India | SFO | 9 | No | Omicron | BA.2 |
| 1/22/2022 | South Africa | SFO | 10 | Yes | Omicron | BA.1 |
| 1/22/2022 | India | SFO | 10 | No | Omicron | BA.2 |
| 1/23/2022 | Germany | EWR | 15 | No | Omicron | BA.1 |
| 1/23/2022 | South Africa | EWR | 12 | Yes | Omicron | BA.1.1 |
| 1/23/2022 | India | EWR | 15 | No | Omicron | BA.2 |
| 1/23/2022 | South Africa | SFO | 10 | Yes | N/A | ND |
| 1/23/2022 | Germany | SFO | 10 | N/A | N/A | ND |
| 1/23/2022 | South Africa | SFO | 5 | Yes | N/A | ND |

Abbreviations: ATL, Hartsfield-Jackson Atlanta International Airport; EWR, Newark Liberty International Airport; JFK, John F. Kennedy International Airport; SFO, San Francisco International Airport; SGTF, S-Gene Target Failure; VOC, Variants of concern; WHO, World Health Organization
<sup>a</sup>N/A - not applicable indicates samples were not run on a RT-PCR assay that would result in SGTF for any known SARS-CoV-2 variants
<sup>b</sup>ND - variant lineage and/or sub-lineage not determined

## Supplementary Methods

### Enrollment Process

Passengers and flight crew arriving from pre-selected international destinations were invited to participate in the voluntary program at one of four US international airports. During September 29–November 27, flight crew announced the program during selected international flights from India using a provided script and directed passengers to informational brochures. During November 28-January 23, in-flight announcements were not used as pooled samples were grouped by country of origin rather than flight. During both enrollment periods, program information and booths were placed in a strategic location at each airport, in the arrivals area near baggage claim, to engage interested arriving travelers. Guidance for collection of dry nasal swab and saliva self-collection samples were provided in technical instructions [1-2].

We estimated the total number of eligible participants on pre-selected international flights using the following methodology: for a sample week, we reviewed the flight numbers indicated on the program pamphlet by the participants and estimated the total number of eligible passengers assuming ∼200 passengers per flight.

### Viral Whole Genome Sequencing

Reverse transcribed RNA was amplified using the ARTICv3 protocol [3]. Amplicons were pooled and prepared for library preparation using standard protocols. For Illumina sequencing, samples underwent tagmentation and were sequenced on NovaSeq 6000 (2×50 bp; Illumina). For rapid lineage identification, a ligation-based library was prepared and sequenced on GridION (Oxford Nanopore). With SARS-CoV-2 genome MN908947.3 as reference, Illumina data was aligned with BWA-MEM and Guppy-called Nanopore data with minimap2 (using ont-map option). Variant Call Format (VCF) files were generated with FreeBayes [4] (Illumina) or medaka-variant (Nanopore) and these VCF files used in both cases to generate consensus sequences with bcftools. Lineages were called from consensus sequences with pangolin v3.1.16 [5] using the sarscov2_lineages workflow on the Terra platform [6]. Variant lineages were not assigned for sequences that did not meet pangolin minimum length of 10,000 bases and <50% N (unassigned) bases. Clade assignments, reference-based QC calls, mutational profiles, and associated visualizations for analyses were produced through the Nextclade webserver v1.8.1 [7]. The presence or absence of minor variant strains was assessed through analysis of alternate allele frequencies in the Illumina data set generated by both FreeBayes (v1.3.1) and iVar (v1.3.1). For mixed samples, lineage deconvolution was performed using two independent workflows: LCS (https://arxiv.org/abs/2110.01117v1) and Freyja (https://www.medrxiv.org/content/10.1101/2021.12.21.21268143v1.full.pdf).

## Footnotes

1 See e.g., 45 C.F.R. § 46.102(l)(2); 21 C.F.R. part 56; 42 U.S.C. §241(d); 5 U.S.C. §552a; 44 U.S.C. §3501 et seq

## References

1. Centers for Disease Control and Prevention COVID-19 Response Team. SARS-CoV-2 B. 529 (Omicron) Variant—United States, December 1–8, 2021. MMWR Morbidity and mortality weekly report 2021; 70(50): 1731–4.

2. Lambrou AS, Shirk P, Steele MK, et al. Genomic Surveillance for SARS-CoV-2 Variants: Predominance of the Delta (B. 1.617. 2) and Omicron (B. 1.1. 529) Variants—United States, June 2021–January 2022. Morbidity and Mortality Weekly Report 2022; 71(6): 206.

3. Thermo Fisher Scientific Confirms Detection of SARS-CoV-2 in Samples Containing the Omicron Variant with its TaqPath COVID-19 Tests. Available at: https://thermofisher.mediaroom.com/2021-11-29-Thermo-Fisher-Scientific-Confirms-Detection-of-SARS-CoV-2-in-Samples-Containing-the-Omicron-Variant-with-its-TaqPath-COVID-19-Tests. Accessed 12/21/2021.

4. Tyson JR, James P, Stoddart D, et al. Improvements to the ARTIC multiplex PCR method for SARS-CoV-2 genome sequencing using nanopore. BioRxiv 2020.

5. Prevention CfDCa. How to Report COVID-19 Laboratory Data. Available at: https://www.cdc.gov/coronavirus/2019-ncov/lab/reporting-lab-data.html#what-to-report. Accessed December 28, 2021.

6. Ginkgo Bioworks. Concentric by Ginkgo and XpresCheck™ Confirm First North American Detections of Novel BA.3 Sublineage of Omicron Variant through CDC COVID-19 Air Travel Biosecurity Program. Available at: https://www.prnewswire.com/news-releases/concentric-by-ginkgo-and-xprescheck-confirm-first-north-american-detections-of-novel-ba3-sublineage-of-omicron-variant-through-cdc-covid-19-air-travel-biosecurity-program-301447761.html. Accessed 3/4/2022.

7. Johansson MA, Wolford H, Paul P, et al. Reducing travel-related SARS-CoV-2 transmission with layered mitigation measures: symptom monitoring, quarantine, and testing. BMC medicine 2021; 19(1): 1–13.

8. Swadi T, Geoghegan JL, Devine T, et al. Genomic evidence of in-flight transmission of SARS-CoV-2 despite predeparture testing. Emerging infectious diseases 2021; 27(3): 687.

9. Khanh NC, Thai PQ, Quach H-L, et al. Transmission of SARS-CoV 2 during long-haul flight. Emerging infectious diseases 2020; 26(11): 2617.

10. Hahn SM, McMeekin JA. Coronavirus (COVID-19) Update: FDA Alerts Consumers About Unauthorized Fraudulent COVID-19 Test Kits. US Food and Drug Administration 2020.

11. Abdalhamid B, Bilder CR, McCutchen EL, Hinrichs SH, Koepsell SA, Iwen PC. Assessment of specimen pooling to conserve SARS CoV-2 testing resources. American journal of clinical pathology 2020; 153(6): 715–8.

12. Hamer DH, Rizwan A, Freedman DO, Kozarsky P, Libman M. GeoSentinel: past, present and future. Journal of Travel Medicine 2020; 27(8): taaa219.

## Supplemental References

1. Instructions for Supervised Pooled Testing Available at: https://support.concentricbyginkgo.com/hc/en-us/articles/360059837572-Instructions-for-Supervising-Pooled-Testing. Accessed 1/5/2022

2. Patient Self-Collection Kit and Collection Process Standard Operating Procedures. Accessed December 30, 2021. https://www.fda.gov/media/137782/download

3. Tyson JR, James P, Stoddart D, et al. Improvements to the ARTIC multiplex PCR method for SARS-CoV-2 genome sequencing using nanopore. Preprint. bioRxiv. 2020;2020.09.04.283077. Published 2020 Sep 4. doi:10.1101/2020.09.04.283077

4. Garrison E, Marth G. Haplotype-based variant detection from short-read sequencing. arXiv preprint 1207.3907 2012

5. Áine O’Toole, Emily Scher, Anthony Underwood, Ben Jackson, Verity Hill, John T McCrone, Rachel Colquhoun, Chris Ruis, Khalil Abu-Dahab, Ben Taylor, Corin Yeats, Louis du Plessis, Daniel Maloney, Nathan Medd, Stephen W Attwood, David M Aanensen, Edward C Holmes, Oliver G Pybus, Andrew Rambaut, Assignment of epidemiological lineages in an emerging pandemic using the pangolin tool, Virus Evolution, Volume 7, Issue 2, December 2021, veab064, https://doi.org/10.1093/ve/veab064

6. https://app.terra.bio/

7. ksamentov, I., Roemer, C., Hodcroft, E. B., & Neher, R. A., (2021). Nextclade: clade assignment, mutation calling and quality control for viral genomes. Journal of Open Source Software, 6(67), 3773, https://doi.org/10.21105/joss.03773

8. Global initiative on sharing avian flu data (GISAID) database. Accessed December 21, 2021. https://www.gisaid.org/

